# What positives can be taken from the COVID-19 pandemic in Australia?

**DOI:** 10.1101/2020.12.10.20247346

**Authors:** S Cornell, B Nickel, E Cvejic, C Bonner, KJ McCaffery, J Ayre, T Copp, C Batcup, JMJ Isautier, T Dakin, RH Dodd

**Author notes:** **Funding:** This study was not specifically funded, but in-kind support was provided by authors with research fellowships. **Conflict of Interests:** The authors declare that they have no competing interests. **Corresponding author:** Samuel Cornell, School of Public Health. Room 128A Edward Ford Building (A27), The University of Sydney, NSW, 2006.

## Abstract

**Objective:** To investigate whether Australians have experienced any positive effects during the COVID-19 pandemic, despite the disruption to society and daily life.

**Methods:** National online longitudinal survey. As part of a June 2020 survey, participants (n=1370) were asked ‘In your life, have you experienced any positive effects from the COVID-19 pandemic’ (yes/no), with a free-text explanation if yes, and also completed the WHO-Five well-being index. Differences were explored by demographic variables. Free-text responses were thematically coded.

**Results:** 960 participants (70%) reported experiencing at least one positive effect during the COVID-19 pandemic. Living with others (p=.045) and employment situation (p<.001) at baseline (April), were associated with experiencing positive effects. Individuals working for pay from home were more likely to experience positive effects compared to those who were not working for pay (aOR=0.45, 95%CI: 0.32, 0.63, p<.001), or who were working for pay outside the home (aOR=0.40, 95%CI: 0.28, 0.58, p<.001). Age and education were not associated with positive effects when controlling for employment and household numbers. There was an overall effect of gender (p=.001), where those identifying as female were more likely than males (aOR=1.62, 95%CI: 1.25, 2.09) to report experiencing a positive effect. 54.2% of participants reported a sufficient level of wellbeing, 23.2% low wellbeing and a further 22.6% very low wellbeing. Of those experiencing positives, 945/960 (98%) provided an explanation. The three most common themes were ‘Family time’ (33%), ‘Work flexibility’ (29%), and ‘Calmer life’ (19%).

**Conclusion:** A large proportion of surveyed Australians reported positive effects resulting from changes to daily life due to the COVID-19 pandemic in Australia. Enhancing these aspects may build community resilience to cope with future pandemic responses. The needs of people living alone, and of those having to work outside the home or who are unemployed, should be considered by health policy makers and employers in future pandemic preparedness efforts, as these groups were least likely to report positive experiences and may be more vulnerable.

## Introduction

A substantial human toll has resulted from the COVID-19 global pandemic, with over 1.5 million lives lost (1,2) and trillions cost to the global economy (3). Nevertheless, the detrimental effects of the pandemic have differed considerably between countries, with different government responses and public health orders implemented. In 2020 Australia has fared favourably in comparison to many other developed nations after closing international borders, intensive COVID-19 testing and contact tracing, in addition to other methods of slowing the spread of the virus (4,5). Early in the course of the pandemic, Australia ceased all inbound travel except for exceptional circumstances and to allow citizens and permanent residents home (6). At this time, citizens and permanent residents were also prohibited from leaving the country (7,8).

This is not the first pandemic or large-scale crisis to disrupt daily life, that humans have experienced. It is however, the first at this scale that has occurred during a time of global connectivity via the internet, telecommunications and air travel (9). Throughout our history as a species, humans have endured famines, plagues, world wars, climate changes, nuclear catastrophes and other near-misses of existential threat (10). In fact, there is widespread perception that the rate of natural disasters is only increasing (11). Exploring how humans may find positives among these demanding circumstances and how collective resilience enables this, may help us mitigate the negative consequences of COVID-19 and future global crises.

Previous research has demonstrated that people can react positively to large scale crises by developing resilience, particularly as a community. This positive reaction to disaster has been observed amongst other populations under duress in times of crises, such as Londoners during The Blitz (12), New Zealanders in the immediate aftermath of the Canterbury Earthquakes (13), and Chileans in the aftermath of the 2010 earthquake and tsunami (14,15) which include reports of community cohesion, a positive outlook and demonstrable traits of resilience. Furthermore, research found that characteristics of community resilience, including tight bonds and a sense of kinship, were helpful in addressing the Ebola virus in Liberia (16).

In this paper we investigated whether participants had experienced any positive effects during the pandemic and what those positives were; and explored whether there were any sociodemographic factors associated with a more or less positive experience during this period.

## Methods

### Study design and setting

The Sydney Health Literacy Lab (SHeLL) has been conducting a national longitudinal survey in Australia since April 2020. The original sample was recruited via an online market research panel, Dynata, and using paid advertising on social media (n=4326). Participants were aged 18 years and over, could read and understand English and were currently residing in Australia. Participants recruited through social media (n=2006) were then followed-up monthly from April-July. Participants recruited via social media were given the opportunity to enter a prize draw for the chance to win one of ten AUD$20 gift cards upon completion of each survey. More details on recruitment are provided elsewhere (17), other survey results are provided elsewhere (17–20). In the June survey (June 5^th^-12^th^), participants were asked the following question, ‘In your life, have you experienced any positive effects from the COVID-19 pandemic’ (yes/no). Those participants responding ‘yes’ were asked to provide a free-text response: ‘Please describe what these positive experiences have been’. Participants also completed the WHO-Five well-being index (WHO-5); a 5-item questionnaire that measures current mental well-being over the previous two weeks (21). We used the STROBE cross sectional checklist when writing our report (22).

### Ethical Approval

This study was approved by The University of Sydney Human Research Ethics Committee (2020/212). All participants in the study provided informed consent before completing the online survey.

### Quantitative analysis

Quantitative data were analysed using Stata/IC v16.1 (StataCorp, College Station TX, USA). Descriptive statistics were generated for demographic characteristics of the analysed sample. Logistic regression was applied to determine whether age (categorised into 10 year groups until 70+), gender (male, female, other/prefer not to say), highest level of educational attainment (high school or less, trade certificate, university education), household structure (live alone, or live with 1-2, 3-4, or 5 or more others) or employment situation in April (not working for pay, working for pay from home, working for pay outside the home, or other working for pay situation) were associated with self-reported positive experiences during the COVID-19 pandemic. Multivariable linear regression was also applied to determine whether the aforementioned variables were associated with participants’ WHO-5 score (scored 0-100), with scores of ≤28 representing very low wellbeing, ≤50 low wellbeing, >50 high wellbeing.

### Content Analysis

Free-text responses were analysed using content analysis (23), a widely used analysis method which combines qualitative and quantitative methods to analyse text data, allowing the content and frequency of categories to be reported. One member of the research team (SC) first read through all the free-text responses and developed the initial coding framework. All members of the research team also reviewed the free-text responses and discussed the coding framework. A random selection (randomised in excel) of 200 responses (∼20%) were double coded independently by two members of the research team (SC and RD). Level of agreement was tested using Cohen’s kappa (24) and indicated substantial agreement (κ=0.83). Any discrepancies were discussed between SC and RD until consensus was reached. SC then coded the remaining 745 responses. The frequency of each code and main themes were then reported.

## Results

### Descriptive statistics

Demographic characteristics of the sample overall, and by their response to the question “In your life, have you experienced any positive effects from the COVID-19 pandemic” are provided in Table 1. Of the 1370 individuals in the sample, 960 (70.1%) indicated that they had experienced at least one positive during the COVID-19 pandemic. Overall, 54.2% (n=743) of participants reported a sufficient level of wellbeing (>50/100), while 23.2% (n=318) showed low wellbeing (≤50/100) and a further 22.6% (n=309) showed very low wellbeing (≤28/100).

**Table 1.**
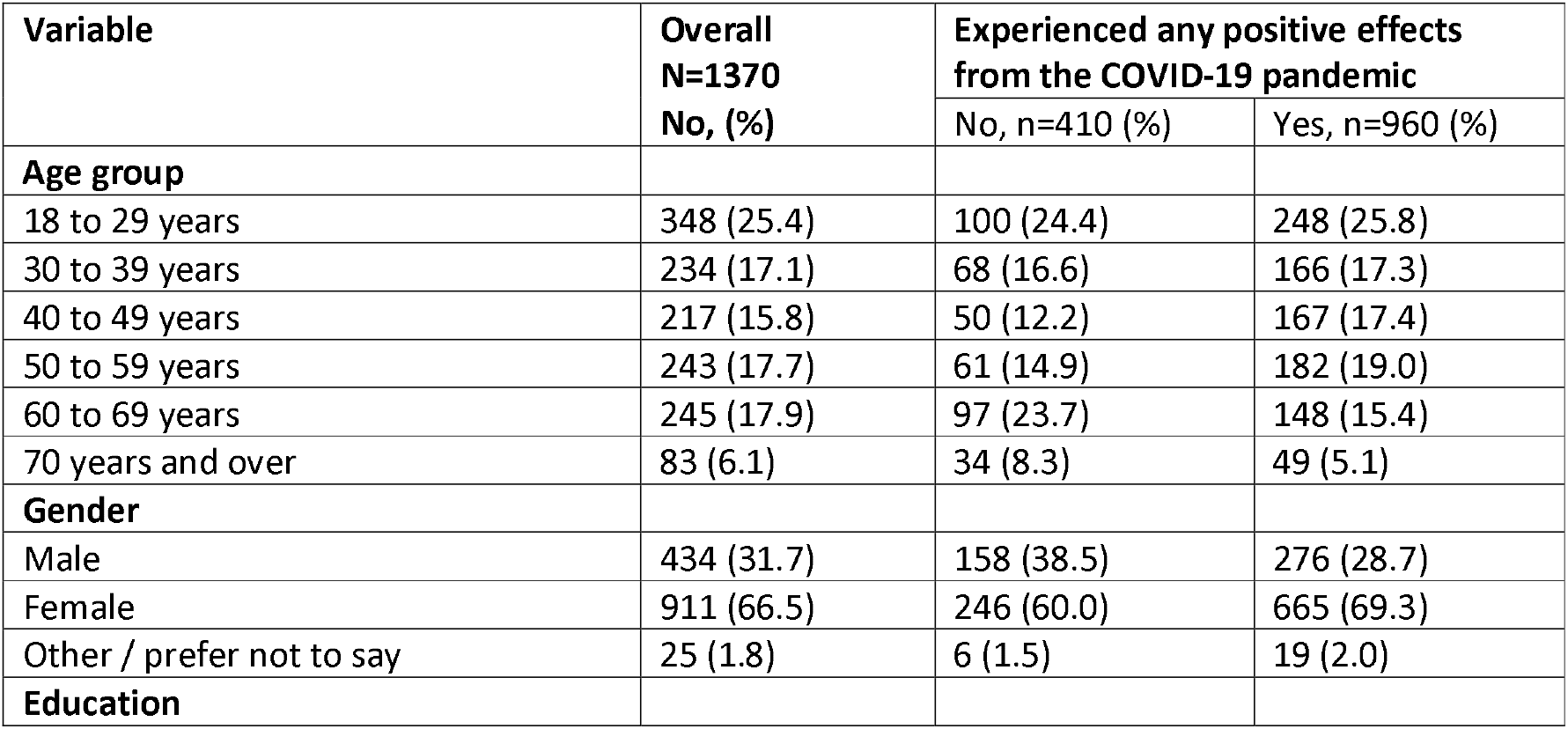
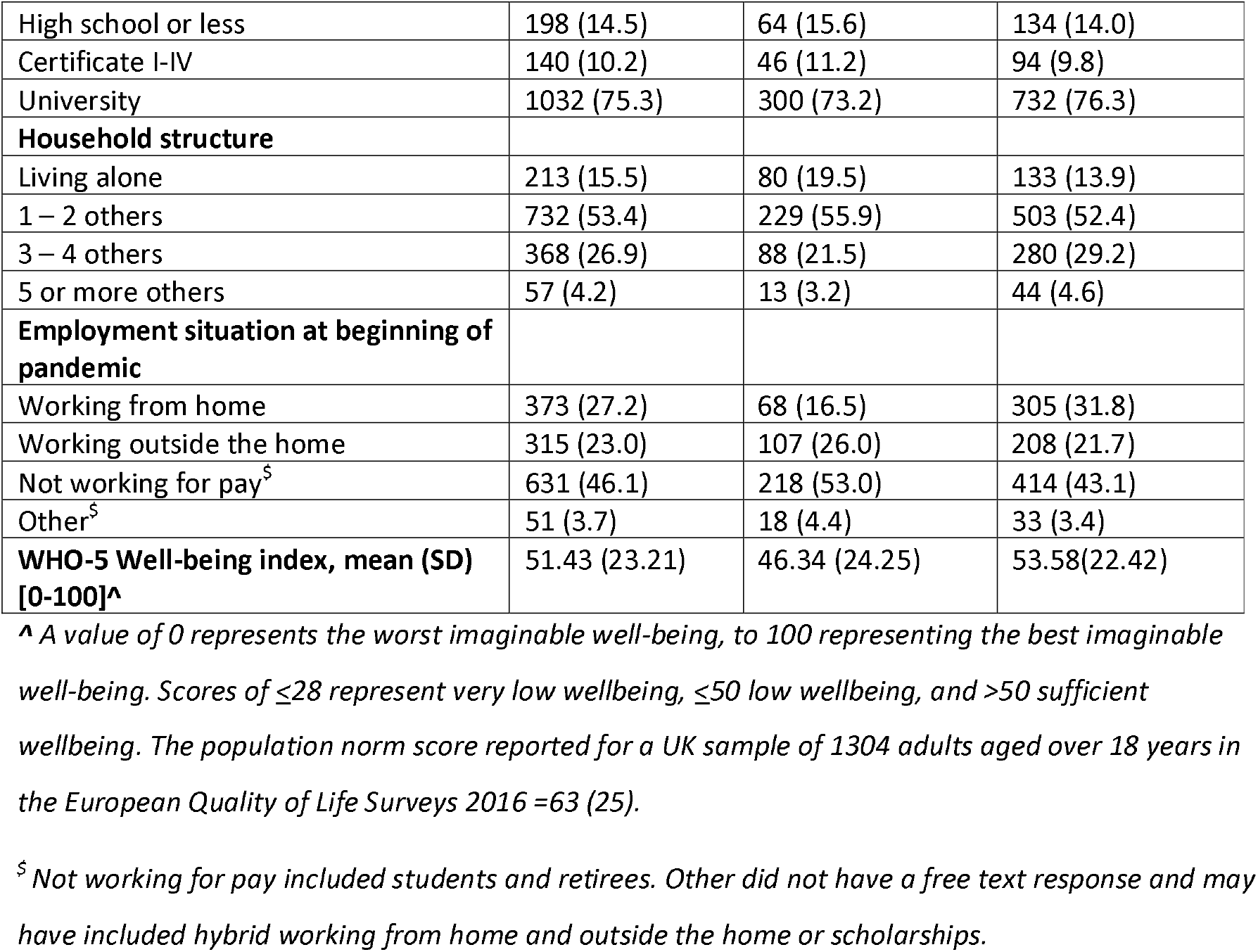
Demographic characteristics of the analysis sample (N=1370). Data are presented as n (%) unless otherwise indicated.

An independent samples t-test indicated that those who reported experiencing any positive effects from the COVID-19 pandemic also had higher wellbeing scores than those who did not report positive effects (mean difference [MD]: 7.25, 95% CI: 4.59, 9.91; t(1369)=5.35, p<.001; Cohen’s d=0.31)

### Factors associated with a positive effect of the COVID-19 pandemic

Adjusted odds ratios from logistic regression are displayed in Table 2. There was an overall effect of gender (p=.001), where those identifying as female were more likely than males (aOR=1.62, 95%CI: 1.25, 2.09, p<.001) to report experiencing a positive effect of the pandemic. Individuals who lived in households with a greater number of people were more likely to experience positive effects (p=.045). Compared to those living alone, individuals who reported living with 3 to 4 others (aOR=1.65, 95%CI: 1.11, 2.45, p=.012) or 5 or more other people (aOR=2.08, 95%CI: 1.03, 4.20, p=.043) had greater odds of reporting a positive effect. Employment situation during the baseline survey (April) was also associated with the experience of positive effects (p<.001); individuals who were not working for pay (aOR=0.45, 95%CI: 0.32, 0.63, p<.001), or who were working for pay outside of the home (aOR=0.40, 95%CI: 0.28, 0.58, p<.001) were less likely to experience positive effects in comparison to those who were working for pay from home. Age and education did not appear to be associated with reporting positive effects of the COVID-19 pandemic when controlling for other model factors including household numbers and employment.

**Table 2.** Results from multivariable logistic regression on the experience of positive effects from the COVID-19 pandemic. Data are presented as adjusted odds ratios (95% confidence intervals).

| Variable | Adjusted OR | 95% CI | p-value |
| --- | --- | --- | --- |
| **Age group** |  |  | .14 |
| 18 to 29 years | Reference |  |  |
| 30 to 39 years | 0.83 | 0.56, 1.23 |  |
| 40 to 49 years | 1.13 | 0.75, 1.72 |  |
| 50 to 59 years | 1.13 | 0.76, 1.66 |  |
| 60 to 69 years | 0.68 | 0.47, 1.00 |  |
| 70 years and over | 0.73 | 0.43, 1.25 |  |
| **Gender** |  |  | <.001 |
| Male | Reference |  |  |
| Female | 1.62 | 1.25, 2.09 |  |
| Other / prefer not to say | 1.90 | 0.73, 4.96 |  |
| **Education** |  |  | .99 |
| High school or less | 1.01 | 0.70, 1.44 |  |
| Certificate I-IV | 0.98 | 0.66, 1.45 |  |
| University | Reference |  |  |
| **Household structure** |  |  | .045 |
| Living alone | Reference |  |  |
| 1 – 2 others | 1.27 | 0.92, 1.77 |  |
| 3 – 4 others | 1.65 | 1.11, 2.45 |  |
| 5 or more others | 2.08 | 1.03, 4.20 |  |
| **Employment situation at beginning of pandemic** |  |  | <.001 |
| Working from home | Reference |  |  |
| Working outside the home | 0.40 | 0.28, 0.58 |  |
| Not working for pay | 0.45 | 0.32, 0.63 |  |
| Other | 0.40 | 0.21, 0.76 |  |

A multivariable linear regression on the WHO-5 well-being index, displayed in table 3, found that participants who were older (50-60, 60-70 and 70+ years) had higher wellbeing than participants in the 18-30 year group (all p<.001). Males had slightly higher wellbeing than females (MD=3.06, 95%CI: 0.44, 5.67, p=.022)and participants with certificate I-IV education (MD=-5.14, 95%CI: −9.14, - 1.13, p=.012), but not those with high school certificate or less (MD=0.72, 95%CI:-2.92, 4.36, p=.70), had lower wellbeing than those who were university educated. Participants who lived alone were found to have lower wellbeing compared to those who lived with 1-2 (MD=4.05, 95% CI: 0.59, 7.50, p=.022), or 3-4 others (MD=7.14, 95%CI: 3.17, 11.11, p<.001). Employment situation was not associated with wellbeing (p=.33).

**Table 3.** Results from multivariable linear regression on WHO-5 well-being index. Data are presented as marginal mean differences (95% confidence intervals) compared to the indicated reference group.

|  | Mean difference | 95% CI | p-value |
| --- | --- | --- | --- |
| **Age group** |  |  | <.001 |
| 18 to 29 years | Reference |  |  |
| 30 to 39 years | -0.23 | -4.12, 3.67 |  |
| 40 to 49 years | 2.11 | -1.86, 6.09 |  |
| 50 to 59 years | 8.14 | 4.34, 11.94 |  |
| 60 to 69 years | 14.47 | 10.51, 18.42 |  |
| 70 years and over | 17.69 | 12.00, 23.38 |  |
| **Gender** |  |  | .019 |
| Male | Reference |  |  |
| Female | -3.06 | -5.67, -0.44 |  |
| Other / prefer not to say | -9.60 | -18.75, -0.44 |  |
| **Education** |  |  | .03 |
| High school or less | 0.72 | -2.93, 4.36 |  |
| Certificate I-IV | -5.14 | -9.14, -1.13 |  |
| University | Reference |  |  |
| **Household structure** |  |  | .006 |
| Living alone | Reference |  |  |
| 1 – 2 others | 4.05 | 0.59, 7.50 |  |
| 3 – 4 others | 7.14 | 3.17, 11.11 |  |
| 5 or more others | 3.69 | -3.00, 10.39 |  |
| **Employment situation at beginning of pandemic** |  |  | .33 |
| Working from home | Reference |  |  |
| Working outside the home | -0.86 | -3.99, 2.28 |  |
| Not working for pay | -0.04 | -3.46, 3.38 |  |
| Other | 5.28 | -1.32, 11.87 |  |

### Content analysis results

Of the 960 participants reporting a positive experience, 945 (98%) provided a written response detailing their positive experiences. 18 themes (plus an ‘other’ category) captured these responses (Table 4).

**Table 4.**
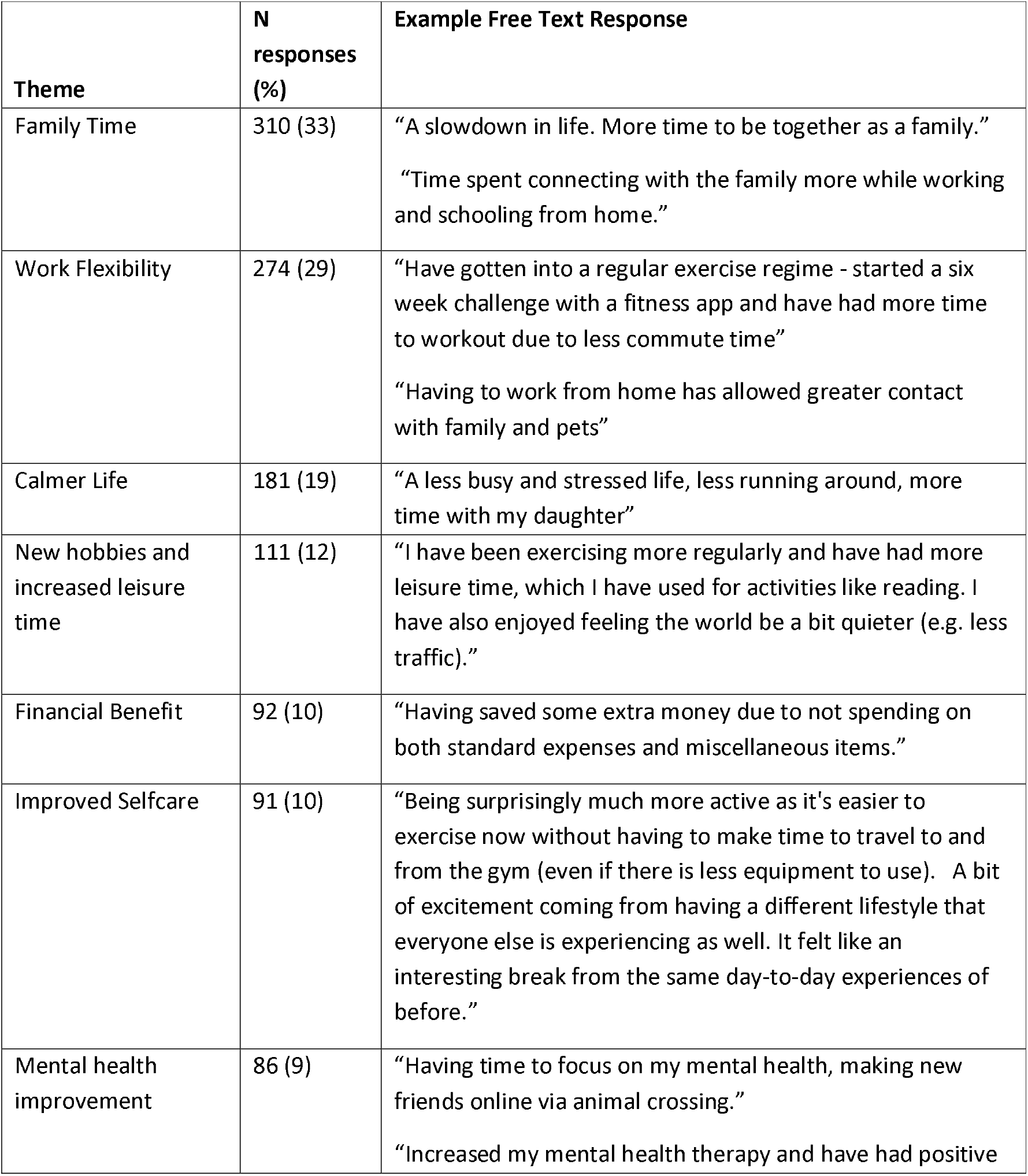
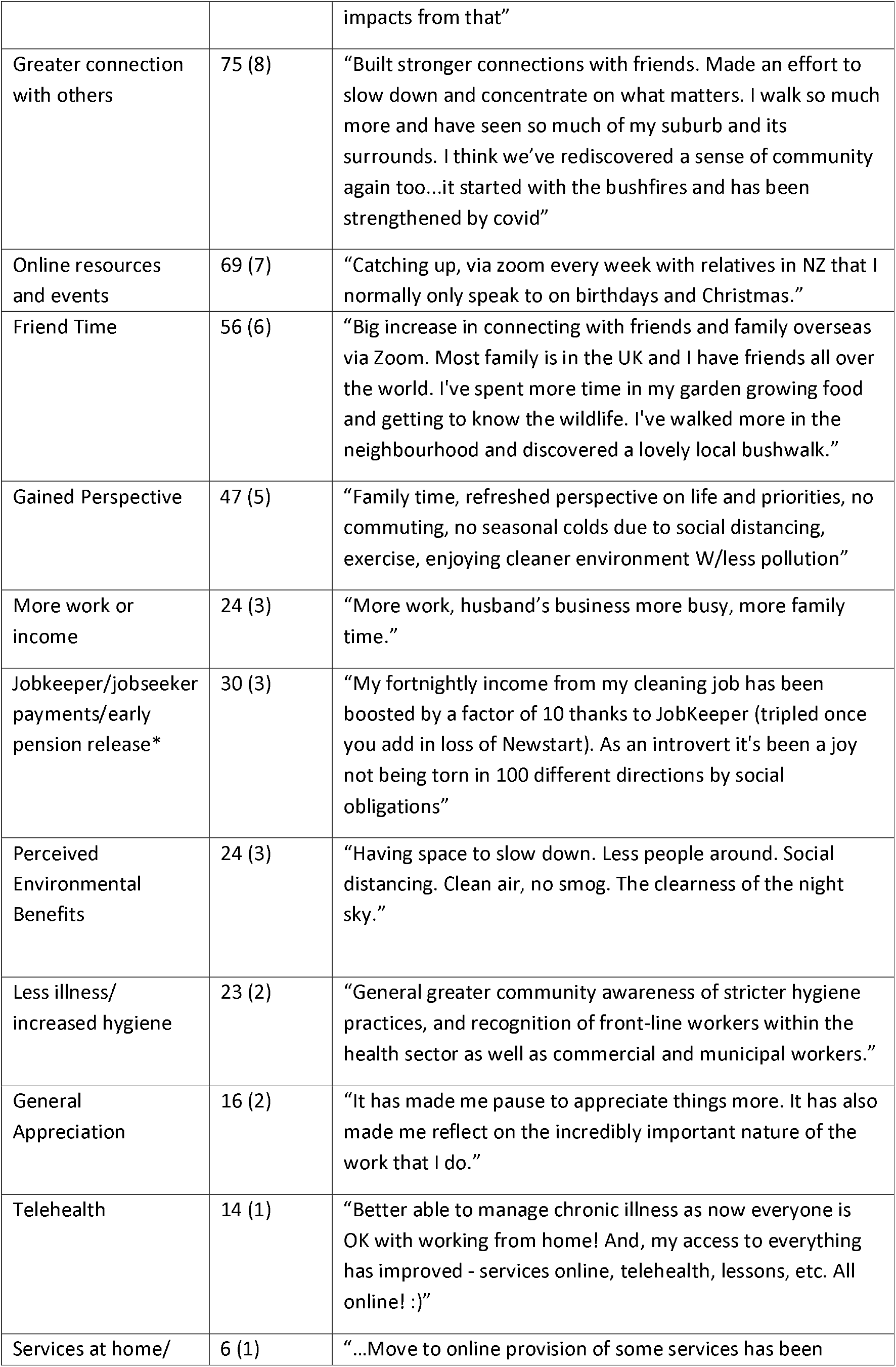
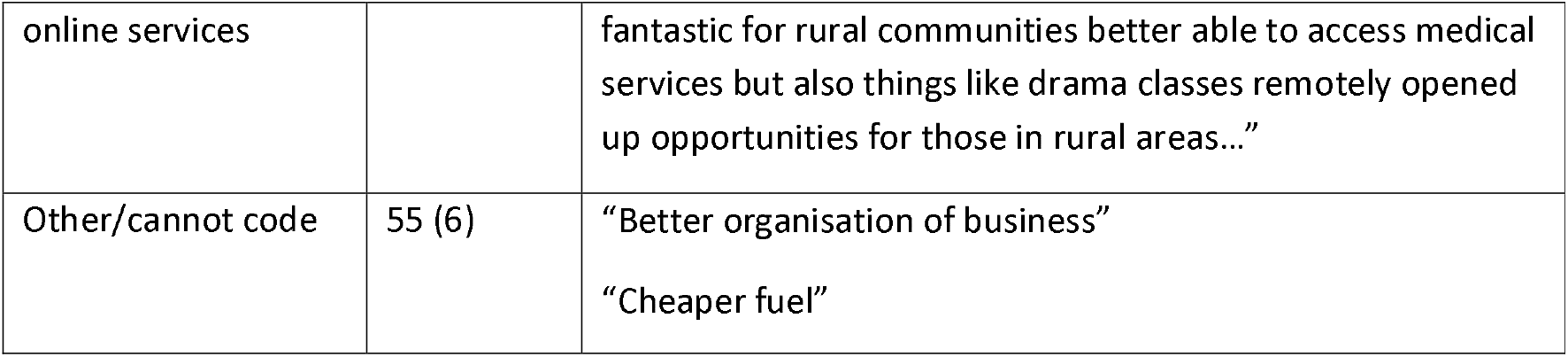
Themes identified in free-text responses to question ‘In your life, have you experienced any positive effects from the COVID-19 pandemic’ with example response.

The three most commonly reported positive impacts identified were;

1. ‘Family Time’ (33%), with participants describing positive effects of being able to have more time with their immediate family and a feeling of greater appreciation for their family members and improvements in their family relationships. Responses to this theme included; *“…allowed my family to get closer together…”* and *“Appreciate close family contact via internet and the company the family I live with provide”*.
2. ‘Work Flexibility’ (29%) with participants discussing an appreciation of increased work flexibility with no commute involved, feeling more productive when they do work and a feeling of being more autonomous and in control of their day. Quotes such as *“No commute time. Usually takes me an hour door to door. It’s been great reclaiming 2 hours per day. It’s a shame my boss wants us to go back to the office now*…*”* and *“Working from home, avoiding commuting and the stresses that can pose in your life, has been a definite positive during COVID-19 isolation and I sincerely hope to strike a balance between office attendance and telecommuting post-COVID. We’ve definitely shown it’s do-able*.*”* Highlighted the connection between working from home and a feeling of empowerment over one’s time.
3. ‘Calmer Life’ (19%) with participants highlighting the stillness of the world around them and showing an appreciation for a less frantic daily life. Quotes to this effect included; *“calm shopping centres, no traffic noises, less trucks, less people parking on street, less places to rush, less crowds”* and *“Everything has been quieter and calmer. Little traffic on roads, shops not as busy. As an introvert, no pressure to join in outings to clubs etc”*.

Other major themes in which over 10% of participants identified positive effects included; 4) Taking up a new hobby/ increase in leisure activity/ time outdoors; 5) Financial benefit/ saving money and 6) Improved selfcare/ exercise/ home cooking.

Themes were often interconnected, with many participants identifying positives that covered several themes. Quotes such as this demonstrate connections between the top themes; *“I am able to work from home full time - that’s 2 hours a day that I’m not wasting commuting. I am loving this. I was able to cycle a lot more when the streets were empty. This was an incredibly positive experience for me. As a woman who is a relatively slow cyclist I am terrified of sharing the road with selfish and angry drivers in huge cars. Therefore it was absolutely freeing and empowering to be able to cycle anywhere and any time and not fear for my life. I have been able to spend more time with my child and be more involved in her education. I have not felt the obligation to catch up with people and my time has been my own. This has been the calmest most productive time of my life by far*.*”*

## Discussion

These findings illustrate that a large proportion of the Australians included in our survey found some positive experiences to take away from the first four months of the COVID-19 pandemic. Of note, a large percentage of participants in this survey found it positive having the opportunity to spend more time with family, appreciated being able to work from home or have more flexibility in their working arrangements, and many described enjoying a less busy lifestyle. However, not all groups were equally likely to experience these positive effects. Those whom were unemployed during our April survey or whom were working for pay outside of the home were less likely to experience positive effects. Those who lived alone were also less likely to experience positives. These groups may need more support for future pandemic restrictions.

It is notable that the predominant theme that was found in the participants’ comments was being able to spend more time with family. Although we acknowledge that many people may have been separated from their families during this time, this sample reported that the lockdown period provided many families a chance to be together and prioritise those relationships. This finding is in keeping with previous research into crises demonstrating that family and community connection is able to attenuate the detrimental impacts of disaster and promote resilience amongst community members (26,27). Furthermore, other research conducted during the pandemic has found a similar effect on increased connection and bonding for families (28). It also indicates a need for greater support for those living alone or away from close family members.

Working from home and workplace flexibility were highly prevalent in the responses from this sample. The pandemic gave many people who traditionally worked regular hours in an office environment a chance to experience a greater amount of freedom, flexibility and autonomy over their work lives. People have been able to save time and money from not commuting, which they have been able to use in other ways. Many people reported feeling more productive and happier with working from home and hoped that it would continue post-pandemic. Previous research has shown that people who have a shorter or no commute tend to be happier than those with a longer commute (29). The pandemic facilitated more people being able to experience a no-commute lifestyle and the benefits this can bring. These changes could be retained after the pandemic response.

Other positives included enjoying a quieter and less busy life. This theme often tied in directly to increased work flexibility and seeing family more. The initial stages of the pandemic in Australia included stay at home orders which varied across states. These periods of time acted as enforced ‘downtime’ for many people whom did not have to leave home to work (28).

It may seem counterintuitive that a wide range of positives were found by surveyed participants during the pandemic, with a large proportion of participants in this study attesting to the ability to spend more time with family and friends, feeling a greater connection to community and enjoying more flexible working practices. However, when considering human adaptation to past crises (30,31), these results are not surprising. Throughout human evolution, people survived and thrived in small groups which were intimate and deeply social (32). Cooperation and reciprocity were key elements to the function of the group. Crises such as the pandemic seem to foster community connection and therefore help to attenuate the negatives of the event (33).

Our survey revealed that people living in single person households were significantly less likely to experience positive effects from the changes to life in the early stages of the pandemic. This finding is important and adds weight to the use of ‘social bubbles’ (designated social and physical interaction between members of different households) to maintain psychological wellbeing for people living alone during the pandemic (34,35).

Although a large proportion of participants in this survey found positives, it is crucial to acknowledge that it is possible to acknowledge positives in a crisis but not necessarily find the overall experience a positive one. Furthermore, many of the reported positives were time-specific and may not have remained as restrictions changed over the course of the pandemic when people returned to the office to work or when they became busy in other areas of their lives again. Furthermore, this research did not include participants from Melbourne post the implementation of the second Victorian lockdown, as this survey was completed in July 2020 before Melbourne re-entered into strict restrictions.

It seems apparent from the results of this study, and in keeping with previous research, that a core tenet of surviving and even thriving through crises is having strong connections with others which facilitates resilience and the ability to find positives or “cheerfulness in the face of adversity” (36). We also need to be aware of groups that cannot respond in such a way, and ensure they receive additional support during future pandemic restrictions.

### Strengths and limitations

This study is novel in its use of both qualitative and quantitative methods to determine if any positive outcomes are to be found in the experiences of a large sample of Australians during the COVID-19 pandemic. The study is limited by its sample which is not nationally representative nor culturally and linguistically diverse. Future research should aim to include a broader representation of experiences of the pandemic most notably those from diverse backgrounds and cultural groups.

## Conclusion

A large proportion of Australians in our survey reported experiencing positive effects because of changes to daily life due to the COVID-19 pandemic in Australia. However, the needs of people living alone or having to work outside the home should be considered by health policy makers and employers in the post-pandemic world as these groups were least likely to experience positive effects. We are lucky to live in a country that has handled the COVID-19 pandemic well overall, not forgetting the extra challenges faced by Victorians and those already experiencing socioeconomic disadvantage or loneliness. By identifying positive experiences that helped people cope with COVID-19 restrictions, we can manage future pandemic responses in ways that promote community resilience. It is important to provide extra support to groups that couldn’t access the benefits of changes to daily life and consider whether we should keep some changes post-pandemic. This might include flexible working and a greater emphasis on local community engagement to promote social connections.

## Data Availability

Data may be accessed upon reasonable request from the authorship team.

## Footnotes

*The Australian government enacted financial policies including ‘Jobkeeper’ (37) and ‘jobseeker’ (38) to support people unemployed during the COVID-19 pandemic and also allowed early access to superannuation (39) (pension) money for individuals whom met certain criteria.

## Contributors

All authors were involved in the conception and design of the study, developing the methods and coordinating the running of the study. SC developed the initial coding framework and all authors contributed to the framework. SC, RD and EC contributed to the analysis. SC drafted the manuscript and RD, BN, EC, CB, KM, JA and TC reviewed and edited the manuscript. All authors have read and agreed to the published version of the manuscript.

## Acknowledgements

We would like to acknowledge and thank the Australian public who participated in this survey.

## Data statement

Data may be accessed upon reasonable request from the authorship team.

